# Individual factors underlie temperature variation in sickness and in health: influence of age, BMI and genetic factors in a multi-cohort study

**DOI:** 10.1101/2021.01.26.21250480

**Authors:** Rose S. Penfold, Maria Beatrice Zazzara, Marc F. Osterdahl, GSTT CovidCollaborative, Carly Welch, Mary Ni Lochlainn, Maxim B. Freidin, Ruth C.E. Bowyer, Ellen Thompson, Michela Antonelli, Yu Xian Rachel Tan, Carole H. Sudre, Marc Modat, Benjamin Murray, Jonathan Wolf, Sebastien Ourselin, Tonny Veenith, Janet M. Lord, Claire J. Steves

## Abstract

**Introduction:** Ageing affects immune function resulting in aberrant fever response to infection. We assess the effects of biological variables on basal temperature and temperature in COVID-19 infection, proposing an updated temperature threshold for older adults.

**Methods:** Participants:

a. <u>Unaffected twin volunteers</u>: 1089 adult TwinsUK participants.
b. <u>London hospitalised COVID-19+</u>: 520 adults with emergency admission.
c. <u>Birmingham hospitalised COVID-19+</u>: 757 adults with emergency admission.
d. <u>Community-based COVID-19+</u>: 3972 adults self-reporting a positive test using the COVID Symptom Study mobile application.

**Analysis:** Heritability assessed using saturated and univariate ACE models; Linear mixed-effect and multivariable linear regression analysing associations between temperature, age, sex and BMI; multivariable logistic regression analysing associations between fever (≥37.8°C) and age; receiver operating characteristic (ROC) analysis to identify temperature threshold for adults ≥ 65 years.

**Results:** Among unaffected volunteers, lower BMI (p=0.001), and older age (p<0.001) associated with lower basal temperature. Basal temperature showed a heritability of 47% (95% Confidence Interval 18-57%).

In COVID-19+ participants, increasing age associated with lower temperatures in cohorts (c) and (d) (p<0.001). For each additional year of age, participants were 1% less likely to demonstrate a fever (OR 0.99; p<0.001).

Combining healthy and COVID-19+ participants, a temperature of 37.4°C in adults ≥65 years had similar sensitivity and specificity to 37.8°C in adults <65 years for discriminating fever in COVID-19.

**Conclusions:** Ageing affects temperature in health and acute infection. Significant heritability indicates biological factors contribute to temperature regulation.

Our observations indicate a lower threshold (37.4°C) should be considered for assessing fever in older adults.

**Key Points:**

- Older adults, particularly those with lower BMI, have a lower basal temperature and a lower temperature in response to infection
- Basal temperature is heritable, suggesting biological factors underlying temperature regulation
- Our findings support a lower temperature threshold of 37.4°C for identifying possible COVID-19 infection in older adults
- This has implications for case detection, surveillance and isolation and could be incorporated into observation assessment

## Background

Normal body temperature of older adults has been observed to be lower than those of younger people, with more limited tolerance of thermal extremes[1,2]. Thermoregulation involves multiple systems, including cardiovascular, respiratory and musculoskeletal. Decreasing basal temperature with age may reflect natural changes in these systems and reduced ability to maintain homeostasis in response to ambient temperature changes.

Normal human body temperature is approximately 37°C (98.6°F); however, variation is observed, with daily variations as much as 0.25 to 0.5°C [3]. Individual biological factors, such as genetics may account for some of the observed variation in human body temperatures. Commercially important species, such as cattle and poultry, show heritability in thermo-regulation under different heat conditions[4,5,6]. However, human heritability data are limited. A small study of 53 adult female twin-pairs (mean age 52) used continuous wrist temperature monitoring to demonstrate Circadian system heritability[7]. Monozygotic pairs showed higher intra-pair correlation than dizygotic twins for most parameters, including temperature, with genetic factors responsible for 46%-70% of variance.

Fever is a physiological response to infection and is clinically important in identifying infection. There is no widespread consensus on the definition of fever, although a number of thresholds have been proposed. Infectious Diseases Society of America guidelines define fever as a single oral temperature >37.8°C (>100°F); repeated oral temperatures >37.2°C (>99°F) or rectal temperatures >37.5°C (>99.5°F); or an increase in temperature of >1.1°C (>2°F) above baseline [8]. Many have questioned applicability of these in older adults [9,10]; indeed, lowering the threshold to 37.2°C has been shown to increase fever detection in older people[8,11]. A previous study found that only 20-30% of patients aged ≥60 with infection present with a temperature of 37.8°C or above [12,13]. Many older adults display atypical presentations, which may be associated with worse outcomes[14]. Age-related changes in immune function may affect pyrogen production and fever response. Altered immune function, or “immunesenescence”, is a well-established feature of physiological ageing[15]. Immunesenescence affects both innate and adaptive arms of the immune system and is thought to be responsible, at least in part, for an increased incidence and severity of infections in older adults[15]. Proposed age-related changes in adaptive immunity include limited diversity in B- and T-cell receptor repertoire, decreased numbers of naive T and B cells and reduced antigen-specific antibody production[16]. Neutrophil function is also compromised by age and frailty and has been proposed as a possible therapeutic target to enhance immune responses in frail, older adults during infection[17].

Coronavirus Disease-19 is an acute respiratory disease caused by severe acute respiratory syndrome coronavirus 2 (SARS-CoV-2). Fever has been cited as a “typical” symptom of COVID-19, with reported prevalence of between 64-98% in hospitalised patients across a number of international studies[18,19,20,21,22]. Current UK public health guidance proposes a temperature ≥37.8°C to define a possible COVID-19 case[23]. As for other infections, atypical presentations of COVID-19 have been documented in older adults, with symptoms including weakness, headache and delirium, often without fever[19,24,25]. In work done by our research group on point-of-care testing for COVID-19, hypothermia (T<36.0°C) was noted to be an early clinical sign in a minority of patients[26].

Demonstrating relationships between age and temperature in a healthy cohort and cohorts with confirmed viral infection is important in understanding effects of ageing on temperature regulation and the fever response to infection. Identifying a more sensitive fever threshold for older adults can ensure that cases of infectious disease, including COVID-19, are detected at an early stage in this vulnerable population. In this study, we first analysed associations between age, sex, body mass index (BMI) and basal temperature and heritability of temperature in a sample of healthy volunteers. We determined if these associations were consistent with those observed during the acute temperature response to infection, in community-based and hospitalised cohorts with COVID-19. We aimed to identify a more appropriate temperature threshold for discriminating COVID-19 infection in adults ≥ 65 years of age than the widely utilised 37.8°C– the temperature proposed by UK public health guidance to screen for COVID-19+ cases[23].

## Methods

### Participants

This study uses distinct cohorts.

a. <u>Unaffected twin volunteers</u>: Measurements were collected between January 2017-April 2018 on 1089 adult twin volunteers, a convenience sample of the TwinsUK registry enrolled at the Department of Twin Research (DTR), King’s College London (KCL)[27]. Demographic information recorded on twins in this cohort includes age, sex, zygosity and BMI. <u>Hospital COVID-19+ cohorts:</u> Data was collected as part of the international COVIDCollab study, led by the Geriatric Medicine Research Collaborative[28,29]. All data is routinely collected and includes age, sex, BMI, and vital signs, including temperature. Full data collection information is published previously[28]. Only cases of COVID-19infection confirmed by RT-PCR of nasopharyngeal swab were included. For patients readmitted during study period, data from index admission was used.
b. <u>London</u>: First documented point-of-care temperature measurements were collected on all adult patients with COVID-19 infection confirmed by RT-PCR of nasopharyngeal swab, with unscheduled admission to St Thomas’ Hospital, London between March 1st-May 4th, 2020;
c. <u>Birmingham</u>: First documented point-of-care temperature measurements were collected on all adult patients with confirmed COVID-19 infection, with unscheduled admission to Queen Elizabeth Hospital, Birmingham, between March 4th, 2020-April 13th, 2020.
d. <u>Community-based “app” COVID + cohort:</u> Data was obtained from UK participants enrolled in the COVID Symptom Study mobile application “app” (KCL & Zoe Global Ltd.). The app captures information related to COVID-19 symptoms self-reported or reported-by-proxy. It was developed by Zoe with scientific input from researchers and clinicians at KCL and was launched in the UK on 24th March 2020. On first accessing the app, users record location, age, ethnicity and health risk factors (supplementary materials). Consent to sharing and analysis of data is obtained during signup for the app. Data was extracted for all UK participants self-reported to have tested positive for COVID-19 between 24^th^ March-9^th^ June 2020 who self-reported “fever or chills” and logged at least one recorded temperature using the app.

### Measurements

(a) <u>Unaffected volunteers</u>: Three infrared tympanic temperature measurements were taken for each twin participant during a single routine research visit to the Guy’s and St Thomas’ Clinical Research Facility. A mean temperature for each participant was used for analysis.
(b) & (c) <u>Hospital COVID-19+ cohorts:</u> Patient records were reviewed. Only COVID-19 cases confirmed by RT-PCR of nasopharyngeal swab were included. First documented oral or tympanic temperature on admission was used to reflect temperature during acute infection.
(d) <u>Community “app” COVID-19+ cohort:</u> Self-reported temperature measurements were extracted for all adult participants self-reported to have tested positive for COVID-19, who had recorded at least one temperature measurement using the app. Maximum temperature was used for analysis, to likely reflect temperature during acute illness.

### Analyses

Descriptive statistics were used to describe demographic and key clinical characteristics of the study cohorts.

Differences between the two hospital COVID-19+ cohorts (b&c) and differences within the unaffected volunteers cohort (a) were compared using the Fisher exact test for categorical and Wilcoxon signed rank for continuous variables.

To analyse associations between age and temperature in the unaffected volunteer cohort (a), a linear mixed effect model was used, with age, gender and BMI set as fixed factors and accounting for family and zygosity as random factors. In hospital COVID-19+ cohorts (b&c), multivariable linear regression was used to model the effects of age, sex and BMI on temperature recorded on admission. In the community-based COVID-19+ cohort (d), multivariable linear regression was used to analyse associations between age, sex and BMI and maximum self-recorded temperature in the context of confirmed COVID-19. For each cohort, two analyses were performed, the first including age and sex as predictor variables, the second including age, sex and BMI. Participants with missing temperature data were excluded from analysis. Participants with missing BMI data were only excluded for analyses using BMI.

In COVID-19+ hospital and community cohorts (b,c&d), logistic regression was performed to ascertain associations between recorded fever (defined as temperature≥ 37.8°C) and age, adjusted for sex and BMI. A complete case approach was used, with low levels of missing data expected.

Receiver Operating Characteristic (ROC) analysis was performed on a combined dataset of unaffected healthy volunteers (a) and hospitalised COVID-19+ participants (b&c) to identify a temperature threshold in older adults (≥ 65 years of age) with similar sensitivity and specificity for discriminating COVID-19 to a temperature ≥37.8°C in adults <65 years of age.

Genetic and environmental contributions of basal temperature in TwinsUK cohort was estimated using a univariate model[30]. The model decomposes the observed variance of the basal temperature into additive genetic variance (A), shared environment variance (C) and non-shared environment variance (E), deducing the genetic influence of a trait and by comparison of the different nested models. The analysis was carried out using Open Mx[30] for R on residuals of linear regression adjusted for age and sex and BMI. Heritability of self-reported fever was carried out in 3099 [291(9.4%) reporting fever] COVID Symptom Study app participants who were also enrolled in TwinsUK, using biometric modelling for the liability threshold model, which, for a categorical trait, assumes an underlying continuous liability that follows a normal distribution[31].

Data analysis and graphics were performed in the R statistical environment (version 4.0) using the Tidyverse[32], Open Mx[30] and lme4[33] packages, in Stata (version 15.0) and in Python (version 3.9).

### Results

Baseline characteristics and temperature measurements for all cohorts are shown in Table 1. 6/520 patients in the London hospital cohort and 80/757 in the Birmingham hospital cohort had missing temperature data and were excluded from analysis. Of included participants, 28/515 in the London hospital cohort and 31/677 in the Birmingham hospital cohort had missing BMI data and were excluded from analyses including BMI.

**Table 1.** Baseline characteristics and temperature measurements of (a) unaffected volunteers; (b) London and (c) Birmingham hospitalised cohorts with PCR-confirmed COVID-19 and (d) Community-based “app” cohort with self-reported confirmed COVID-19. Categorical variables presented as count (%) and continuous variables as mean (standard deviation). All presented as mean/standard deviation. *mean value of 3 recordings of basal tympanic temperature; ** first recorded temperature on hospital admission; *** maximum self-recorded temperature.

|  | **(a) Unaffected**<br>**volunteers**<br>**(n=1089)** | **(b) London**<br>**Hospital**<br>**(n= 514)** | **(c) Birmingham**<br>**Hospital (n=677)** | **(d) Community**<br>**“app” (n=3967)** |
| --- | --- | --- | --- | --- |
|  | *Unaffected*<br>*Volunteers* | *COVID-19 +* | *COVID-19 +* | *COVID-19 +* |
| **Age (years)** | 56.6/16.8 | 61.9/17.1 | 67.8/17.0 | 42.9/12.8 |
| **Sex (females)** | 914 (84%) | 202 (39%) | 321 (43%) | 2770 (70%) |
| **BMI (kg/m<sup>2</sup>)** | 25.8/5.2 | 28.5/6.8<br>(n=486) | 29.2/7.2<br>(n=646) | 28.2/6.2 |
| **Temperature (°C)** | 36.6/0.4* | 37.5/1.1** | 36.9/0.9** | 37.6/1.0*** |

### Regression Analysis

Results of the linear mixed effect model in the TwinsUK sample and linear regression models in the community-based and hospital cohorts are displayed in Table 2. Increasing age is associated with both lower basal temperature, and lower temperature during illness, whilst increasing BMI is associated with higher basal and illness temperature.

**Table 2.** Results from linear mixed effect model for (a) TwinsUK unaffected volunteers; multivariable linear regression for hospitalised cohorts from (b) London and (c) Birmingham with confirmed COVID-19 and (d) Community-based “app” cohort with self-reported confirmed COVID-19. Beta coefficients are reported in bold and standard errors in brackets; P-values are Bonferroni adjusted at 5%.

|  | **(a) TwinsUK Cohort**<br>**(n=1089)** |  | **(b) London Hospital**<br>**(n= 520)** |  | **(c) Birmingham**<br>**Hospital (n=757)** |  | **(c) Community**<br>**“app” (n=3967)** |  |
| --- | --- | --- | --- | --- | --- | --- | --- | --- |
|  | *Unaffected Volunteers* |  | *COVID-19 +* |  | *COVID-19 +* |  | *COVID-19 +* |  |
|  | Basal Temperature |  | First Recorded<br>Temperature on<br>Admission |  | First Recorded<br>Temperature on<br>Admission |  | Maximum Self-Reported<br>Temperature |  |
|  | Model 1<br>(age and<br>sex) | Model 2<br>(age, sex<br>and BMI) | Model 1<br>(age and<br>sex) | Model 2<br>(age, sex<br>and BMI) | Model 1<br>(age and<br>sex) | Model 2<br>(age, sex<br>and BMI) | Model 1<br>(age and<br>sex) | Model 2<br>(age, sex<br>and BMI) |
| **Age** | **-0.214**<br>(0.035)<br>p<0.001 | **-0.240**<br>(0.036)<br>p<0.001 | **-0.007**<br>(0.003)<br>p=0.040 | **-0.005**<br>(0.003)<br>p=0.320 | **-0.007**<br>(0.002)<br>p<0.001 | **-0.006**<br>(0.002)<br>p<0.001 | **-0.006**<br>(0.001)<br>p<0.001 | **-0.006**<br>(0.001)<br>p<0.001 |
| **Sex** | **0.122**<br>(0.035)<br>p<0.001 | **0.128**<br>(0.035)<br>p<0.001 | **-0.028**<br>(0.098)<br>p=1.000 | **-0.016**<br>(0.101)<br>p= 1.000 | **0.064**<br>(0.072)<br>p=1.000 | **0.053**<br>(0.070)<br>p=1.000 | **0.078**<br>(0.034)<br>p=0.043 | **0.086**<br>(0.034)<br>p=0.031 |
| **BMI** | --- | **0.100**<br>(0.032)<br>p=0.001 | --- | **0.0157**<br>(0.007)<br>p= 0.101 | --- | **0.0087**<br>(0.005)<br>p=0.295 | --- | **0.0136**<br>(0.003)<br>p<0.001 |

Logistic regression, exploring the relationship between age and temperature ≥ 37.8°C, demonstrated that for each additional year of age, participants with confirmed COVID-19 were 1% less likely to show a fever in both the app (OR 0.99; P<0.001) and the Birmingham hospital cohorts (OR 0.99; p=0.040) (Table 3).

**Table 3.**
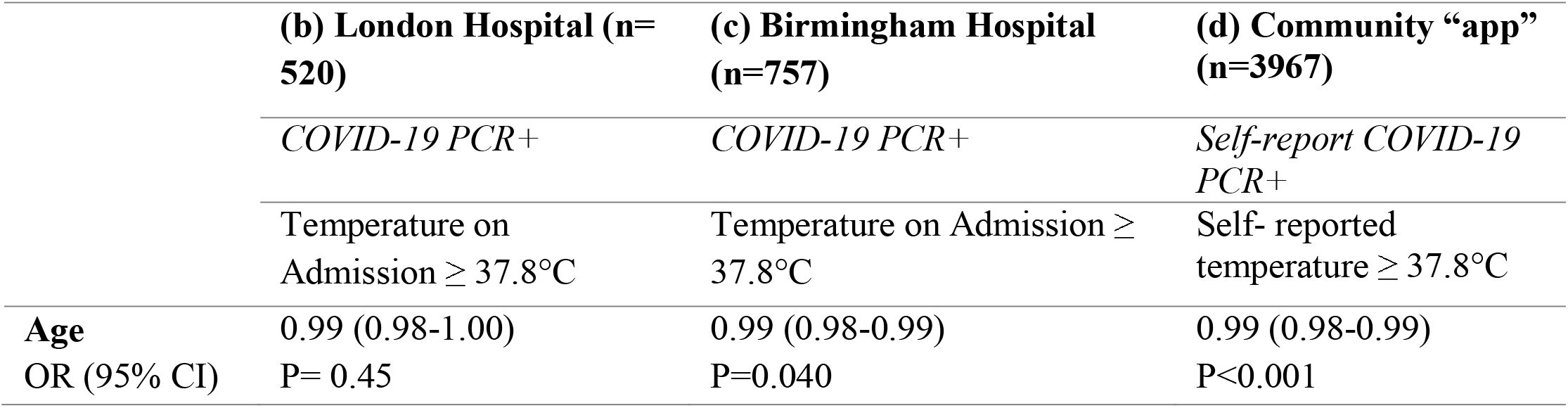
Results of logistic regression for (b) London Hospital cohort and (c) Birmingham Hospital cohort, adjusted for sex and BMI and (d) Community-based “app” cohort. Odds ratios (ORS) are reported in bold, Confidence Intervals (CI) in brackets. Fever is defined as (b) recorded temperature ≥37.8°C and (c) self-reported temperature ≥37.8C.

### ROC analysis

A temperature of 37.8°C had a sensitivity of 33.1% and specificity of 100% for discriminating between the unaffected volunteer cohort and hospitalised COVID-19+ patients in a combined dataset. ROC analysis performed on a combined dataset of unaffected healthy twins volunteers and hospitalised COVID-19+ participants showed that a temperature of 37.4°C in adults ≥65 years of age had similar sensitivity (36.2%) and specificity (98.9%) to a temperature of 37.8°C in adults <65 years of age for discriminating COVID-19+ patients from unaffected volunteers.

**Figure 1.**
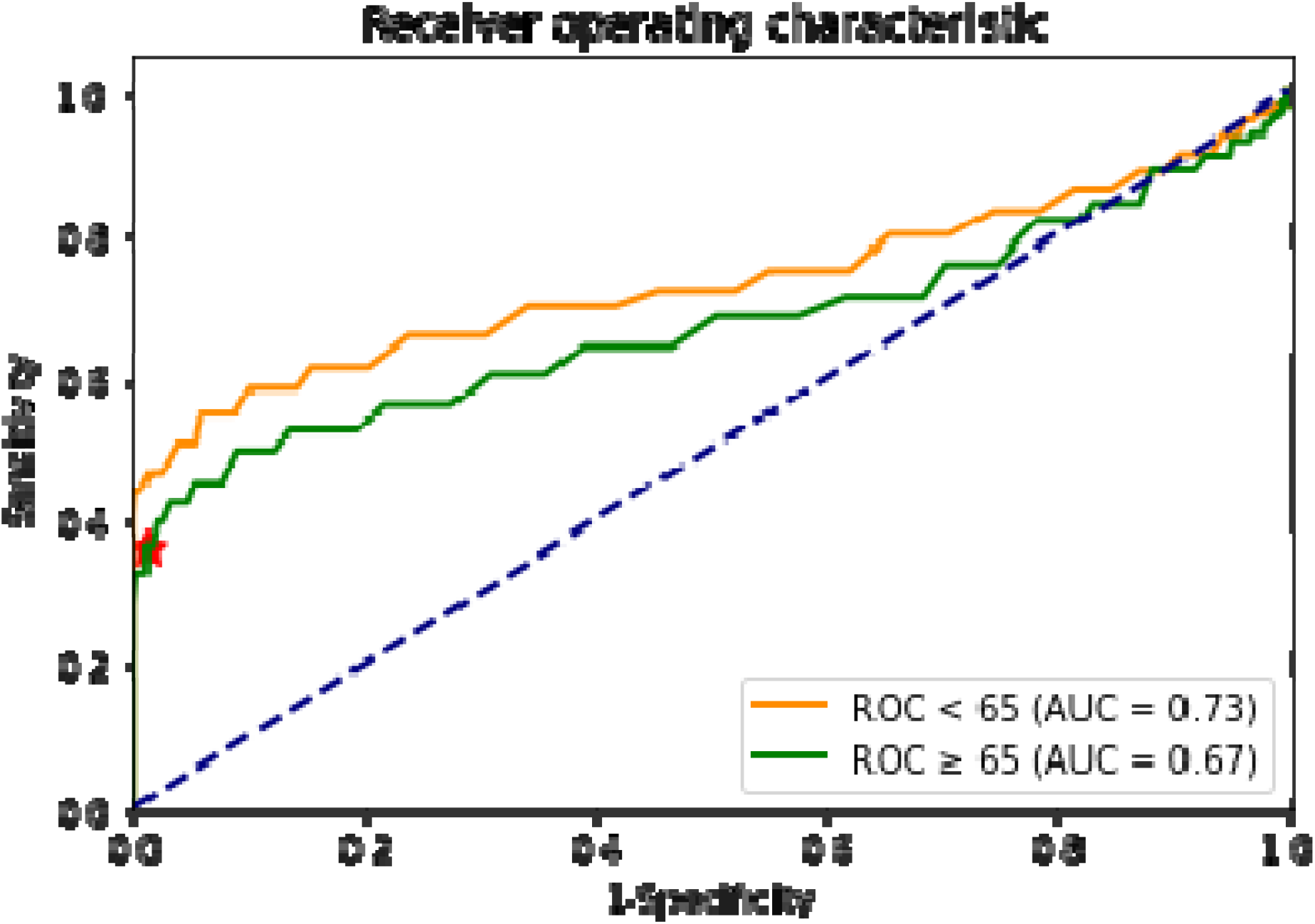
ROC curves in patients <65 years of age (orange) and ≥ 65 years of age (green); the red star indicates the point characterised by a True Positive Rate of 0.36 and False Positive Rate of 0.01.

### Heritability analysis

Results for heritability analysis are summarised in Table 4.

**Table 4:** Results for Heritability Analysis for Basal Temperature adjusted for age and sex; basal temperature adjusted for age, sex and BMI, and Fever.

| Trait | Model | Variance components |  |  |  | Model fit |  |  | Model comparison |  |  |
| --- | --- | --- | --- | --- | --- | --- | --- | --- | --- | --- | --- |
| | | A | C | E | es | −2LL | AIC | $\chi^2$ | df | p | base |
| **Temperature**<br>**(Adjusting for**<br>**Age and Sex)** | Saturated | - | - | - | 10 | 921.88 | -1236.111 | - | - | - | - |
|  | **ACE** | **0.47[0.18-0.57]** | **0.03[0-0.28]** | **0.49[0.42-0.57]** | **4** | **923.73** | **-1246.262** | **1.85** | **6** | **0.933** | **Saturated** |
|  | AE | 0.50[0.43-0.57] | - | 0.49[0.42-0.57] | 3 | 923.79 | -1248.205 | 0.06 | 1 | 0.810 | ACE |
|  | CE | - | 0.42[0.34-0.48] | 0.57[0.51-0.65] | 3 | 934.40 | -1237.598 | 10.61 | 1 | 1.092e-03 | ACE |
|  | E | - | - | 1.00 [1.00-1.00] | 2 | 1033.16 | -1140.835 | 109.37 | 1 | 1.346e-25 | AE |
| **Temperature**<br>**(Adjusting for**<br>**Age, Sex and**<br>**BMI)** | Saturated | - | - | - | 10 | 913.39 | -1244.602 | - | - | - | - |
|  | **ACE** | **0.44[0.15-0.57]** | **0.06[0-0.31]** | **0.49[0.42-0.57]** | **4** | **915.40** | **-1254.594** | **2.01** | **6** | **0.918** | **Saturated** |
|  | AE | 0.50[0.43-0.57] | - | 0.49[0.42-0.57] | 3 | 915.61 | -1256.384 | 0.21 | 1 | 0.646 | ACE |
|  | CE | - | 0.24[0.35-0.49] | 0.57[0.50-0.64] | 3 | 924.83 | -1247.169 | 9.22 | 1 | 2.139e-03 | ACE |
|  | E | - | - | 1.00 [1.00-1.00] | 2 | 1025.86 | -1148.136 | 110.25 | 1 | 8.646e-26 | AE |
| **Fever** | Saturated | - | - | - | 13 | 1853.05 | -4318.95 | - | - | - | - |
|  | ACE | 0 [0-0] | 0.21 [0-0.38] | 0.79 [0.62-0.98] | 8 | 1856.93 | -4329.07 | 3.88 | 7 | 0.793 | Saturated |
|  | AE | 0.2 [0-0.21] | - | 0.8 [0.66-0.99] | 7 | 1858.20 | -4329.80 | 1.27 | 1 | 0.260 | ACE |
|  | **CE** | **-** | **0.21 [0.02-0.38]** | **0.79 [0.62-0.98]** | **7** | **1856.93** | **-4331.07** | **0.00** | **1** | **1.000** | **ACE** |
|  | E | - | - | 1.00 [1.00-1.00] | 6 | 1861.85 | -4328.15 | 4.92 | 1 | 0.027 | CE |

Analysis of monozygotic twin intra-pair correlation showed a clear heritability signal in comparison to dizygotic twin pairs for basal temperature. When adjusted for age and sex, genetic factors accounted for 47% (95% CI = [18-57]%) of variance in mean temperature in the best model (ACE) as compared to the other models (Saturated, ACE, CE, E) (Table 4). When adjusted for age, sex and BMI, genetic factors accounted for 44% (95% CI = [15-57]%) of variance in mean temperature in the best model (ACE)(Table 4).

Heritability of fever, using twins who are App participants, did not demonstrate a genetic component to temperature response to infection, which may be explained by overwhelming environmental factors driving exposure to SARS-CoV-2 (Table 4).

## Discussion

### Sources of variability in temperature: Age, BMI and heritability

In this study, observed associations between age and temperature were replicated across healthy volunteer, hospitalised COVID-19 and community-based COVID-19 cohorts. Temperature regulation is affected by many physiological processes, including vasomotor sweating function, skeletal muscle response, temperature perception, and physical behaviours, many of which have been described in previous studies and reviews[1,34]. These processes change with ageing and our observation that increasing age reduces both basal temperature and temperature in response to SARS-CoV-2 infection supports this. Case reports and small cohort studies have previously shown that older adults, especially those with frailty or multiple chronic conditions, may not display a clinically significant fever response to infection [12,24]. This may reflect both a lower baseline temperature in older adults, as well as a lower temperature change in response to infection. Age-related reductions in pro-inflammatory mediators in response to infection, such as interleukin-1 and tumour necrosis factor, may also have an effect. In older adults, infection should therefore be suspected even in the absence of fever.

The possible association between higher BMI, often considered a pro-inflammatory state, and an increased basal and fever temperature would appear to concord with this finding for age. The observed associations between BMI and temperature in unaffected volunteer and community-based COVID-19 cohorts were largely independent of age. Some earlier studies, often with smaller, specific samples, demonstrated a conflicting association between BMI and temperature [33, 34]. However, our observation is supported by the recent, large CoLaus study of 4224 men and post-menopausal women[35]. Here, other markers of obesity and insulin metabolism were also associated with higher temperature.

In this analysis, adjusted for both age and BMI, we demonstrate a clear heritable component to basal temperature, pointing to other biological variables which may underlie temperature variation. Genetic factors may also play a crucial role in temperature variability among individuals of different ages and with different BMI. Physiological factors related to age and temperature control may also be heritable. In contrast to basal temperature, the trait “fever”, defined here as a temperature of 37.8°C or above, did not demonstrate a significant genetic component, suggesting that environmental factors (in this case exposure to the SARS-CoV-2 virus) may be the main driver of fever within a population.

### Fever Threshold for Discriminating Infection in Older Adults

Our ROC analysis combining unaffected volunteers and hospitalised COVID-19 patients supports observations from previous studies and concerns from frontline clinicians that current fever thresholds may be inappropriate for older adults[8,11]. A lower temperature of 37.4°C had a similar sensitivity for discriminating the COVID-19 cohorts in adults ≥ 65 years of age as the widely utilised threshold of 37.8°C in younger adults. Our observation is supported by a previous study that demonstrated optimised sensitivity and specificity thresholds of 37.9°C for older adults and 37.3°C for younger adults for discriminating influenza infection[36]. Our finding has important utility for screening and management in clinical settings, with a lower threshold in older adults to promptly detect infections and implement containment measures.

### Implications and generalisability

Associations observed in this study have important clinical implications for case detection and surveillance of infectious diseases, including COVID-19. There is a risk of cases being missed in older adults if inappropriately high fever thresholds are used as a screening or diagnostic tool for COVID-19 infection. This may be compounded in those with lower BMI, for example as a result of sarcopenia, who have lower temperatures. Our findings support use of a lower temperature threshold for older adults to initiate appropriate testing and isolation precautions in hospital settings. This may be relevant for care homes and long-term care facilities, with older populations vulnerable to outbreaks of respiratory disease and high fatality rates. Observations from our community-based cohort may have relevance when considering methods for community case detection and limitation of spread (such as quarantining or screening for travel).

### Strengths and Limitations

Our study combines three types of data: a classic research cohort (TwinsUK), routinely collected clinical data from hospital cohorts, and a cohort of community ‘citizen science’ data. The TwinsUK cohort benefits from a large number of observations, collected using rigorous study procedures. Reassuringly, associations observed in this cohort were recapitulated in the “real-life” hospital and community-based data. This concordance increases generalisability from the academic sphere to clinical and community-based settings.

For historical reasons, the TwinsUK cohort is predominantly female. Females generally show greater individual temperature variability than males. The app cohort was also predominantly female. However, both hospital cohorts were predominantly male, reflecting that men are more severely affected by COVID-19. However, our models did not show any association with sex and recorded temperature, suggesting that these differences in cohort structure would not affect results.

Although the app cohort only contained people with self-reported COVID-19 testing status, it is possible that maximal temperature logged in the app was not synchronous with COVID-19 infection or could be the result of another infection. Only those reporting “fever or chills” were prompted to enter a temperature measurement. Some may not have logged on the day of their maximal temperature. App users did under-represent older age groups (proportion ≥65 years 11.7%, compared to 18.3% of the UK population [37]. 4.8% of app users with a confirmed positive COVID-19 test were ≥65 years of age, which may reflect shielding in this group. This is supported by the fact that ONS data shows lower rates of antibody seropositivity in the older, non-patient facing population [38]. Sampling using a mobile app will under-represent individuals without mobile devices, including those from more deprived backgrounds and marginalised groups, at greater risk for severe disease. Furthermore, testing in the community was not universally available during the period of data collection. The method for temperature was not documented, although oral thermometers are most widely available in the UK. These limitations would be most likely diminish power to detect effects but would not be likely to alter the key association seen between age and temperature which is replicated across cohorts.

Hospitalised patients present at different stages of illness; hence, temperature on admission will not always capture the febrile component of infection. Some patients may have received antibiotics, antipyretics or corticosteroids prior to initial temperature recording. BMI data was missing for a small proportion (<5%) of patients - likely not missing at random, with difficulty gaining accurate heights and weights for example due to extremes of body habitus or severe frailty.

## Conclusion

The significant associations observed between age and basal temperature are important in highlighting effects of ageing on thermoregulation and the immune response. The observed heritability of basal temperature in TwinsUK unaffected participants suggests that biological factors underlie differences in temperature regulation which require further elucidation. As well as demonstrating lower temperatures in older adults, we report lower temperature with decreasing BMI. These two factors may compound each other in older, frail sarcopenic patients.

Our findings strongly support a lower temperature threshold for identifying possible COVID-19 infection in older adults of 37.4°C. This has important implications for case detection, surveillance and isolation and could be incorporated into observation assessments for older people.

## Data Availability

TwinsUK data used in this study is available upon reasonable request to TwinsUK.
COVIDCollab data are available upon application to the Geriatric Medicine Research Collaborative https://www.gemresearchuk.com/.
App data used in this study are available to bona fide researchers through UK Health Data Research using the following link https://healthdatagateway.org/detail/9b604483-9cdc-41b2-b82c-14ee3dd705f6

## Ethics

TwinsUK main ethics was reviewed and approved by the NHS London – London Bridge Research Ethics Committee (REC reference EC/04/015), and by Guy’s and St Thomas’ NHS Foundation Trust Research and Development (R&D) in 2012. TwinsUK BioBank was approved by NHS North West - Liverpool East Research Ethics Committee (REC reference 19/NW/0187), IRAS ID 258513.

The Covid Symptom Study App Ethics has been approved by KCL ethics Committee REMAS ID 18210, review reference LRS-19/20-18210. All subscribers provided consent when signing up for the app.

COVIDCollab project service evaluation approved by Guy’s and St Thomas’ NHS Trust audit leads. Reference number 10777.

The University Hospitals Birmingham (UHB) validation was performed as part of a service evaluation with a (CARMS-16005), after approval on routinely collected anonymised dataset.

## Data sharing

TwinsUK data used in this study is available upon reasonable request to TwinsUK.

COVIDCollab data are available upon application to the Geriatric Medicine Research Collaborative https://www.gemresearchuk.com/.

App data used in this study are available to bona fide researchers through UK Health Data Research using the following link https://healthdatagateway.org/detail/9b604483-9cdc-41b2-b82c-14ee3dd705f6

## Funding and Acknowledgements

The app was developed by Zoe Global Limited with input from King’s College London (KLC) and Massachusetts General Hospital. Support for this study was provided by the NIHR-funded Biomedical Research Centre based at GSTT NHS Foundation Trust. In the funding section: Zoe provided in kind support for all aspects of building, running, and supporting the app and service to all users worldwide.

Investigators also received support from the Wellcome Trust, the <u>Medical Research Council</u> (MRC), BHF, Europena Union (EU), <u>National Institute for Health Research (NIHR)</u>, <u>Chronic Disease Research Foundation (CDRF)</u>, and the NIHR-funded BioResource, Alzheimer’s Society, Clinical Research Facility and <u>Biomedical Research Centre</u> (BRC) based at GSTT NHS Foundation Trust in partnership with KCL, NIHR Birmingham BRC, a partnership between University Hospital Birmingham NHS Trust and University of Birmingham. MNL is supported by an NIHR Doctoral Fellowship (NIHR300159). The views expressed are those of the authors and not necessarily those of the Department of Health and Social Care.

TwinsUK is funded by the Wellcome Trust, MRC, EU, CDRF, Zoe Global Ltd,the NIHR-funded BioResource, Clinical Research Facility and BRC based at Guy’s and St Thomas’ NHS Foundation Trust in partnership with KCL.

We would like to acknowledge the following GSTT COVIDCollab collaborators, responsible for London hospital data collection: Rishi Iyer, Rachael Anders, Lindsay Hennah, Gitanjali Amaratunga, Abigail Hobill, Cassandra Fairhead, Amybel Taylor, Henry Maynard, Marc Osterdahl, Maria Dias, Taha Amir, Natalie Yeo, Jamie Mawhinney, Hamilton Morrin, Li Kok, Luca Scott, Aiden Haslam, Gavriella Levinson, Stephanie Mulhern, Stephanie Worrall, Thurkka Rajeswaran, Katherine Stamboullouian, Sophie McLachlan, Karla Griffith, Daniel Muller, Alice O’ Doherty, Baguiasri Mandane, Irem Islek, Alexander Emery, John Millwood-Hargrave, Andra Caracostea, Laura Bremner, Arjun Desai, Aneliya Kuzeva, Carolyn Akladious, Mettha Wimalasundera, Mairead Kelly, Sally Aziz, Sinead O’Dwyer, Rupini Perinpanathan, Anna Barnard, Nicole Hrouda, Ismini Panayotidis, Nirali Desai, Hannah Gerretson, Rebecca Lau, Zaynub Ghufoor, Hanna Nguyen, Torben Heinsohn, Jack Cullen, Eleanor Watkins, Vaishali Vyas, Daniel Curley, Niamh Cunningham, Vittoria Vergani, Kelvin Miu, Jack Stewart, Nicola Kelly, Lara Howells, Benyamin Deldar, Ross Sayers, Gracie Fisk, Sri Sivarajan, Tahmina Razzak, Helen Ye, Samiullah Dost, Nikhita Dattani, Catherine Wilcock, Gabriel Lee, Jodie Acott, Hannah Bridgwater, Antia Fernandez, Hesham Khalid, Katherine Hopkinson, Deirdre Green, Hejab Butt, Ayushi Gupta, Madeleine Garner, Hazel Sanghvi, Madeleine Daly, Emily Ross-Skinner, Shefali Patel, Danielle Lis. (All affiliated to Guy’s and St Thomas’ Hospital NHS Foundation Trust).

Written permission obtained to include the names of all individuals listed.

